# Predicting public take-up of digital contact tracing during the COVID-19 crisis: Results of a national survey

**DOI:** 10.1101/2020.08.26.20182386

**Authors:** Young Ern Saw, Edina YQ Tan, Jessica S Liu, Jean CJ Liu

## Abstract

**Background:** In the global outbreak of coronavirus disease 2019 (COVID-19), new digital solutions have been developed for infection control. In particular, contact tracing mobile applications provide a means for governments to manage both health and economic concerns. However, public reception of these applications is paramount to success, and global take-up rates have been low.

**Objective:** In this study, we sought to identify sociodemographic factors predicting voluntary downloads of a contact tracing mobile application.

**Methods:** A sample of 505 adults from the general community completed an online survey. As the primary outcome measure, participants indicated whether they had downloaded a contact tracing application introduced at the national level (“TraceTogether”). As predictor variables, we assessed: (1) participant demographics; (2) behavioral changes on account of the pandemic; and (3) pandemic severity (the number of cases and lockdown status).

**Results:** Within our dataset, the strongest predictor of digital contact tracing take-up was the extent to which individuals had already adjusted their lifestyles because of the pandemic (*Z* = 13.97, *p* < .001). Network analyses revealed that take-up was most related to: using hand sanitizers, avoiding public transport, and preferring outdoor over indoor venues during the pandemic. However, demographic and situational characteristics did not significantly predict application downloads.

**Conclusions:** Efforts to introduce contact tracing applications could capitalize on pandemic-related behavioral adjustments that individuals have made. Given that critical mass is needed for contact tracing to be effective, we urge further research to understand how citizens respond to contact tracing applications.

**Trial Registration:** ClinicalTrials.gov NCT04468581

## 1. Introduction

In May 2020, Google and Apple released the “Exposure Notification System” - an Application Programming Interface that logs: who a phone user has been in contact with, for how long, and at what distance [1]. This release came two months after the coronavirus disease 2019 (COVID-19) outbreak was categorized as a pandemic [2], allowing governments to identify and isolate contacts of confirmed cases through a process known as “contact tracing” [3,4].

Less than a year after the first reported cases, over 33 million individuals have tested positive for COVID-19 worldwide, and more than 1 million have died [5]. To contain the spread, over half the global population has been subjected to lock-downs involving school closures, workplace shutdowns, and/or movement restrictions [6]. Although these lockdowns are effective in tapering the epidemic curve [7], they are costly to the global economy and are unsustainable [8]. On the flipside, however, allowing the virus to spread unhindered can lead to an overwhelmed healthcare system and the severe loss of lives [9,10].

To address both infection control and economic concerns, several countries have turned to contact tracing to keep the economy open [11,12]. Epidemiological modelling suggests that if: (i) cases are effectively identified (through rigorous testing protocols), (ii) contact tracing is comprehensive (identifying all possible exposure), and (iii) contacts are quarantined in a timely manner, this strategy can curb the spread of the virus [4,11]. In the optimal scenario, 80% of contacts are traced on the same day an individual tests positive [3,11].

### 1.1. Conventional versus Digital Contact Tracing

Early in the pandemic (and in previous infectious disease outbreaks), contact tracing was manual [13]. Using a range of interview and surveillance techniques, a human contact tracer would typically identify an average of 36 contacts for each positive case [14]. Although this strategy allows high levels of case detection when there are few cases [15], its labor-intensive format – requiring ∼12 hours of tracing for each positive case [16] – is difficult to scale up. Additionally, individuals who test positive may forget who they have been in contact with, undermining the effectiveness of the process [11].

Given these limitations of manual contact tracing, several mobile phone applications have been developed to allow automation – for example, COVID-Watch in the United States [17], COVIDSafe in Australia [18], and Corona-Warn-App in Germany [19]. These applications primarily track Bluetooth signals from phones in the vicinity [3], capturing contacts without the restraints of staffing or recall biases [4,11]. Further, phone applications can notify individuals swiftly after a contact tests positive, allowing them to be quickly isolated [3].

### 1.2. Understanding Predictors of Take-Up

Despite the potential of digital contact tracing, a recent meta-analysis concluded that given implementation barriers, manual contact tracing should remain the order of the day [12]. One major barrier pertains to the take-up of mobile phone applications. Several modelling studies have assessed parameters needed for the COVID-19 reproduction number *R*_*0*_ to fall below one [3,11,20]. As *R*_*0*_ refers to the number of infections caused by one positive case, a value less than one indicates that the virus has been contained. For this to be achieved, contact tracing applications would need to be downloaded by at least 56% of the population [20] – much higher than the average rate of downloads globally (9%) [21].

To increase uptake, the Gulf state Qatar made it compulsory for residents to use the official contact tracing application [22]. Although this legislation led to high download rates (>90%, [23]), the potential backlash from the public (e.g., because of privacy concerns [24, 25]) means that few countries are likely to follow suit. Correspondingly, public health agencies would benefit from an understanding of what predicts voluntary downloads [26], providing an empirical base to nudge citizens to opt in [27].

### 1.3. The Current Study

Given the urgent need to boost contact tracing applications, we report in this paper the first study identifying socio-demographics factors predicting voluntary take-up. Our study was conducted in Singapore, where the world’s first national application ‘TraceTogether’ was launched in March 2020 [28]. TraceTogether uses a centralized approach adopted by several governments [18]: namely, randomly-generated user IDs are created and shared via Bluetooth with phones in close proximity [29]. When individuals test positive for COVID-19, they consent to add both their own user IDs and those of their contacts to a centralized database. This is used to identify matches, and exposure notifications are then sent from the server to close contacts [30, 31]. (As an alternative model, a decentralized approach could be used where both matches and notifications are made through the user’s phone [32]).

As Singapore was the forerunner of this technology, the application has accrued 2.3 million users in half a year - nearly 40% of Singapore’s resident population, or 50% of all smartphone users [33]. Correspondingly, our study represents a “best case scenario” for application uptakes after several months have passed. In terms of the epidemic curve, our study was conducted between April to July 2020, as the country entered and then exited a lockdown (during April-May). This period covered a peak in daily COVID-19 cases (April: >1000/day, or 175 per million population) that gradually tapered over time (July: >100/day, or 17.5 per million population).

## 2. Methods

### 2.1. Study Design and Population

Between 3 April to 17 July 2020, we recruited 505 adults who met the following eligibility criteria: (i) at least 21 years of age, and (ii) had lived in Singapore for a minimum of 2 years. All participants responded to advertisements placed in online community groups (e.g., Facebook or WhatsApp groups for residential estates, universities, and workplaces), or to paid online advertisements targeting Singapore residents.

Prior to study enrolment, participants provided informed consent in accordance with a protocol approved by the Yale-NUS College Ethics Review Committee (#2020-CERC-001; ClinicalTrials.gov registration: NCT04468581). They then completed a 10-minute online survey hosted on the platform Qualtrics [34]. Data collection came from the second phase of a larger study tracking COVID-19 responses, and findings from the first phase have been described elsewhere [35,36].

### 2.2 Outcome Variable: Use of TraceTogether

For the primary outcome variable, participants were asked to indicate if they had downloaded the government’s contact tracing application TraceTogether (binary variable with 1 indicating that they had, and 0 indicating that they had not).

### 2.3 Predictors

#### 2.3.1 Demographics and Situational Variables

As predictors of TraceTogether usage, participants then reported the following demographic data: age (in years), gender, citizenship, ethnicity, marital status, educational level, house type (a proxy of socio-economic status in Singapore), as well as household size. Based on the survey timestamp, we also included as predictors: (i) the total number of cases in Singapore to date, and (ii) whether the nation was in a lock-down at the time of participation (0 = No, 1 = Yes).

#### 2.3.2 Other Behavioral Changes

As a basis of comparison, participants were also asked to identify which of 18 other behavioral changes they had made as a result of the pandemic (apart from downloading the contact tracing application TraceTogether). Specifically, participants were asked whether they had: (1) washed their hands more frequently; (2) used hand sanitizers; (3) worn a mask in public voluntarily (before a law was passed); (4) avoided taking public transport; (5) stayed home more than usual; (6) avoided crowded places; (7) chosen outdoor over indoor venues; (8) missed or postponed social events; (9) changed their travel plans voluntarily; (10) reduced physical contact with others (e.g., by not shaking hands); (11) avoided visiting hospitals and/or healthcare settings; (12) avoided visiting places where COVID-19 cases had been reported; (13) kept a distance from people suspected of recent contact with a COVID-19 case; (14) kept a distance from people who might have recently travelled to countries with an outbreak; (15) kept a distance from people with flu symptoms; (16) relied more on online shopping (e.g., for groceries); (17) stored up more household and/or food supplies than usual; or (18) taken their children out of school. Each item was a binary measure such that 0 = measure was not taken and 1 = measure was taken. These values were then summed to compute an aggregated measure of behavioral change (out of 18), and were included as a predictor to assess whether contact tracing usage was associated with conventional behavioral changes one undertakes during an epidemic [37,38]

As part of the survey, participants were also asked to specify if they had changed their behaviors in any other way (1.8%, *n*=9) or if they had not changed their behaviors at all (0.4%, *n*=2). However, these were removed from statistical analyses due to the low base rate of affirmative responses.

### 2.4. Data Analysis Plan

As the primary analysis, binary logistic regression was used to identify predictors of the contact tracing application TraceTogether. In the first model, participants’ demographics were included as predictors (age, citizenship, gender, marital status, education level, ethnicity, household type, and household size). Citizenship (*base = others*), gender (*base = female*), marital status (*base = single*) and ethnicity (*base = Chinese*) were coded as dummy variables. In the second model, we repeated the first model with the inclusion of situational variables (the total number of COVID-19 cases to date, log-transformed; and lockdown status). Finally, in the third model, we repeated the second model but added the total number of behavioral modifications as a predictor. All data analyses were conducted via the statistical packages SPSS (Ver. 23) and R (3.6.0), with the type 1 decision-wise error rate controlled at α = 0.05.

## 3. Results

### 3.1 Demographics of the Sample

As shown in Table 1, our sample of 505 participants captured a wide range of demographics. Compared to the resident population, the sample was matched in: ethnic composition, household size, and housing type (a proxy of socio-economic status in Singapore) (≤10% difference). However, participants were more likely to be female (62.0 vs. 51.1%), to be single/dating (46.4 vs. 31.6%), to have a higher level of education (12.9 vs. 51.7% without tertiary education), and to be a citizen of Singapore (90.3% vs. 61.4%). Conversely, participants were less likely to subscribe to Buddhism or Taoism (22.3 vs. 43.2%).

**Table 1.**
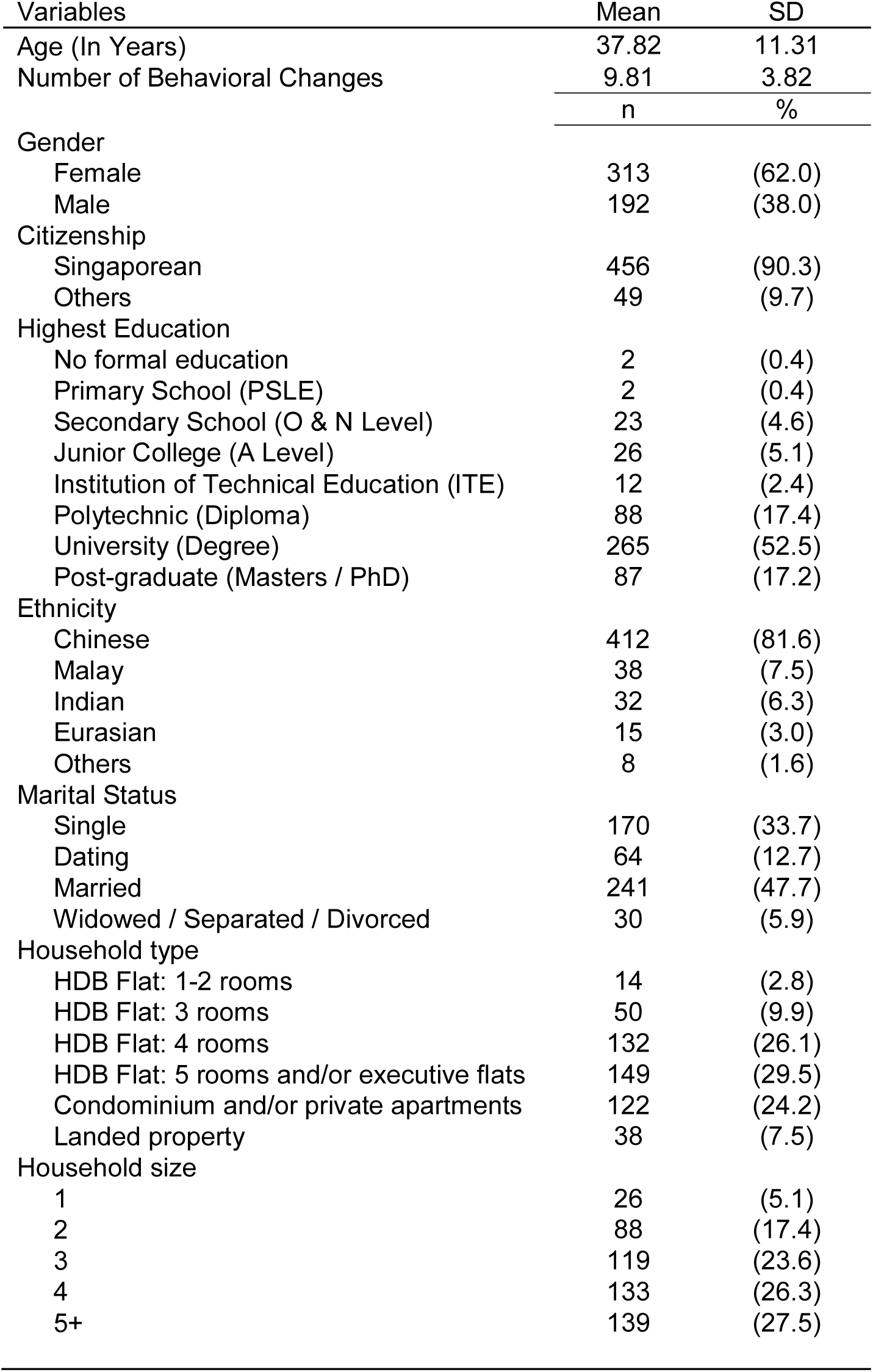
Baseline characteristics of survey respondents (*N* = 505)

### 3.2. Binary Logistic Regression

Of the 505 participants, 274 (54.3%, 95% CI: 49.8% - 58.7%) reported having downloaded TraceTogether, Singapore’s contact tracing application. The download rate in this sample matches that of smartphone users in the resident population [33].

Table 2 shows parameter estimates from logistic regression analyses predicting the use of the application. As can be seen, no demographic or situational variable significantly predicted downloads (Models 1 and 2). After controlling for these variables, the number of behavioral modifications emerged as a significant predictor (Model 3). Namely, with each unit increase in the number of behavioral modifications adopted, participants were 1.11 times more likely to download the contact tracing application (*Z*=13.97, *P*<.001). (For visualization, Table 3 describes the characteristics of users and non-users.)

**Table 2.**
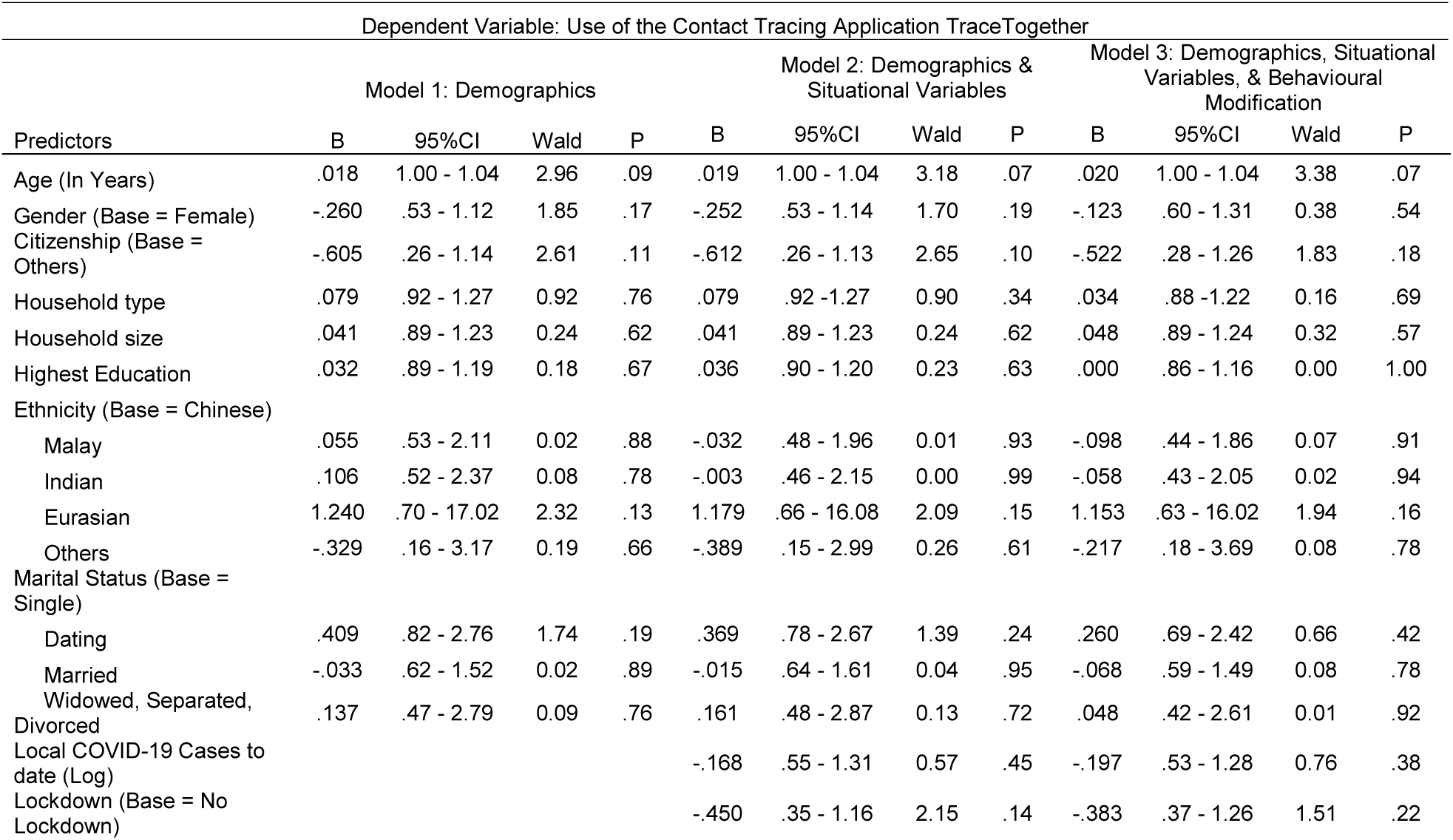
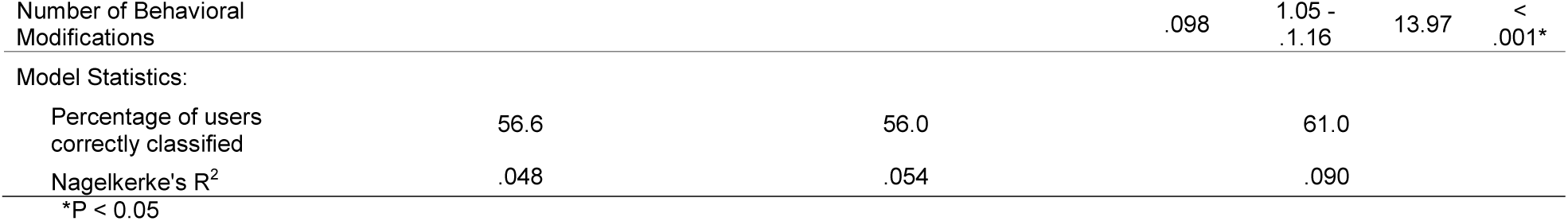
Logistic regression models predicting the use of a contact tracing mobile application

**Table 3.**
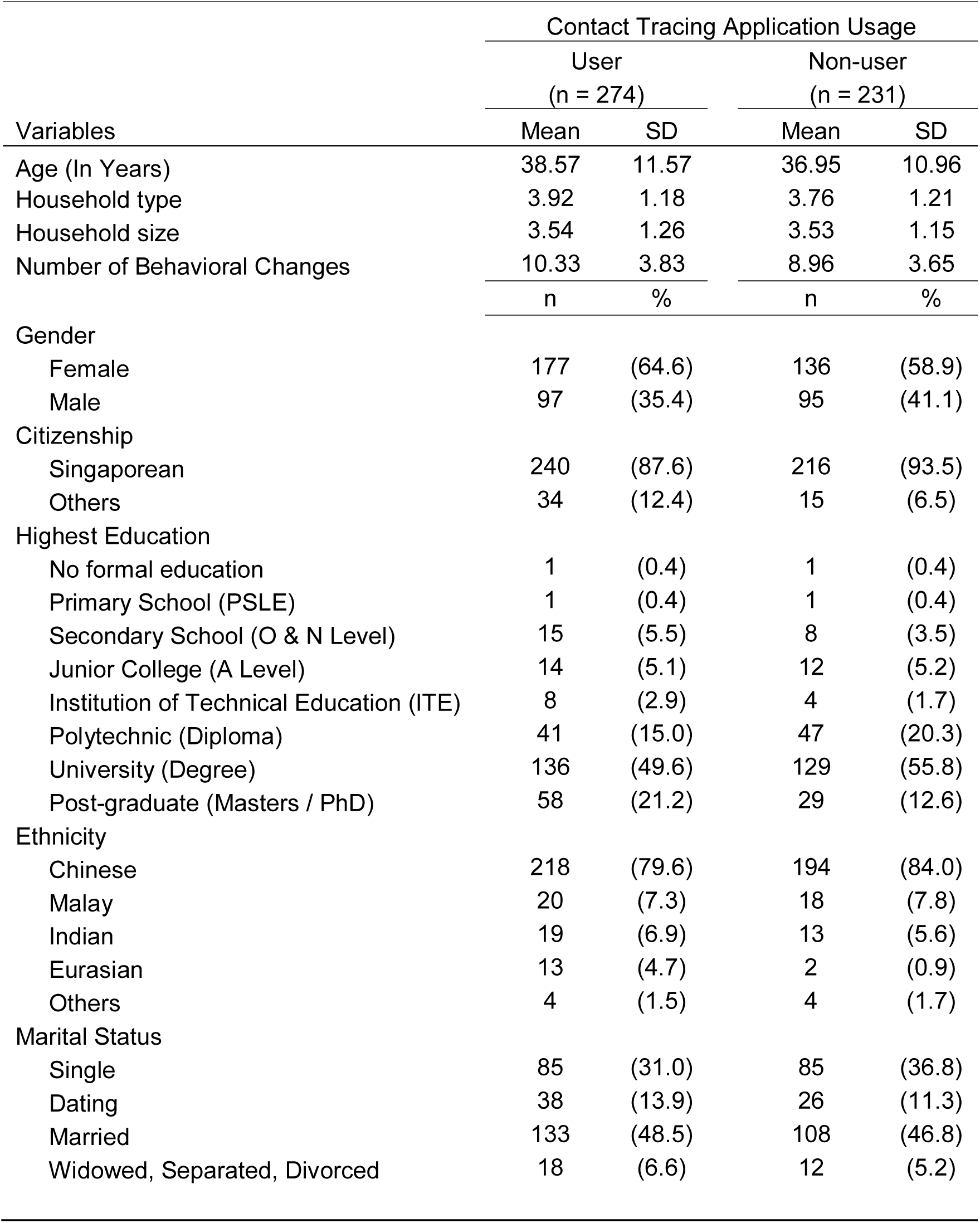
Characteristics of digital contact tracing users

### 3.3 Post-Hoc Network Analysis

In the logistic regression analyses, downloads of the contact tracing application was predicted by the extent to which individuals changed their behaviors because of the pandemic. To understand this association, we conducted further exploratory analyses.

As shown in Figure 1, the majority of participants had modified their behaviors to curb the spread of COVID-19. Use of the contact tracing application (TraceTogether) ranked 10th in the frequency of adoption (54.3%) - approximately equal to voluntary mask-wearing (54.7%).

**Figure 1.**
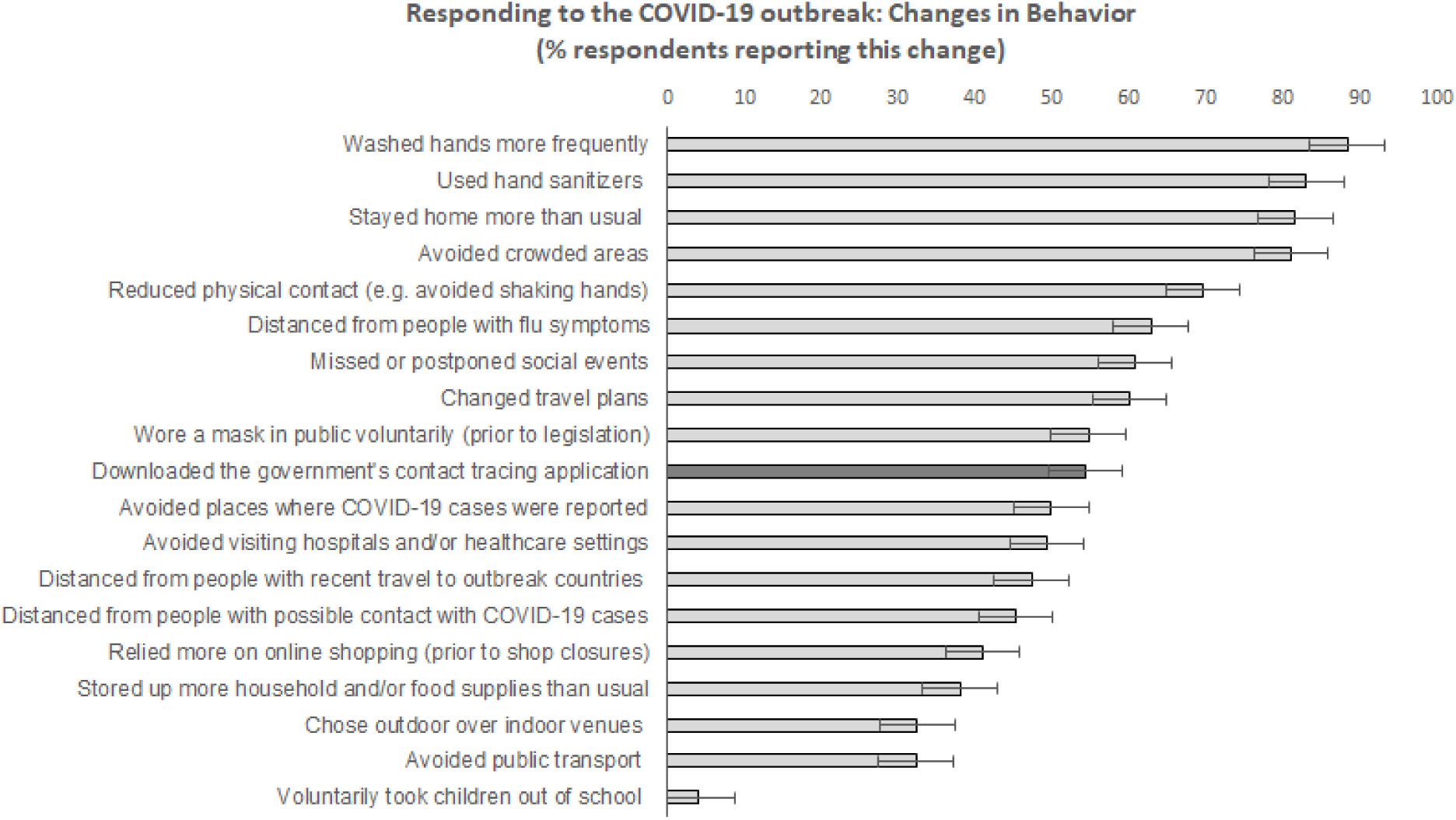
Participants reported what behavioural measures – including the use of digital contact tracing – they had undertaken in response to the COVID-19 outbreak. Horizontal lines represent the 95% confidence intervals.

A corollary question is how the use of digital contact tracing relates to other health protective behaviors - that is, how likely are you to adopt the application if you’ve modified your behaviors in other ways? To address this question, we conducted network analyses by estimating a mixed graphical model (MGM) via the R package *mgm* [39]. MGM constructs weighted and undirected networks where the pathways between the behaviors represent conditionally dependent associations, having controlled for the other associations in the network. Similar to partial correlations, each association (or ‘edge’) is the average regression coefficient of two nodes. To avoid false positive findings, we set small associations to 0 for the main models in the paper.

As shown in Figure 2, use of the contact tracing application was associated with hand sanitizer use, avoiding public transport, and a preference for outdoor vs indoor venues. The adjacency matrix (i.e., numerical values for the average regression coefficient between two nodes) for Figure 2 is presented in Appendix A, Table. S1.

**Figure 2.**
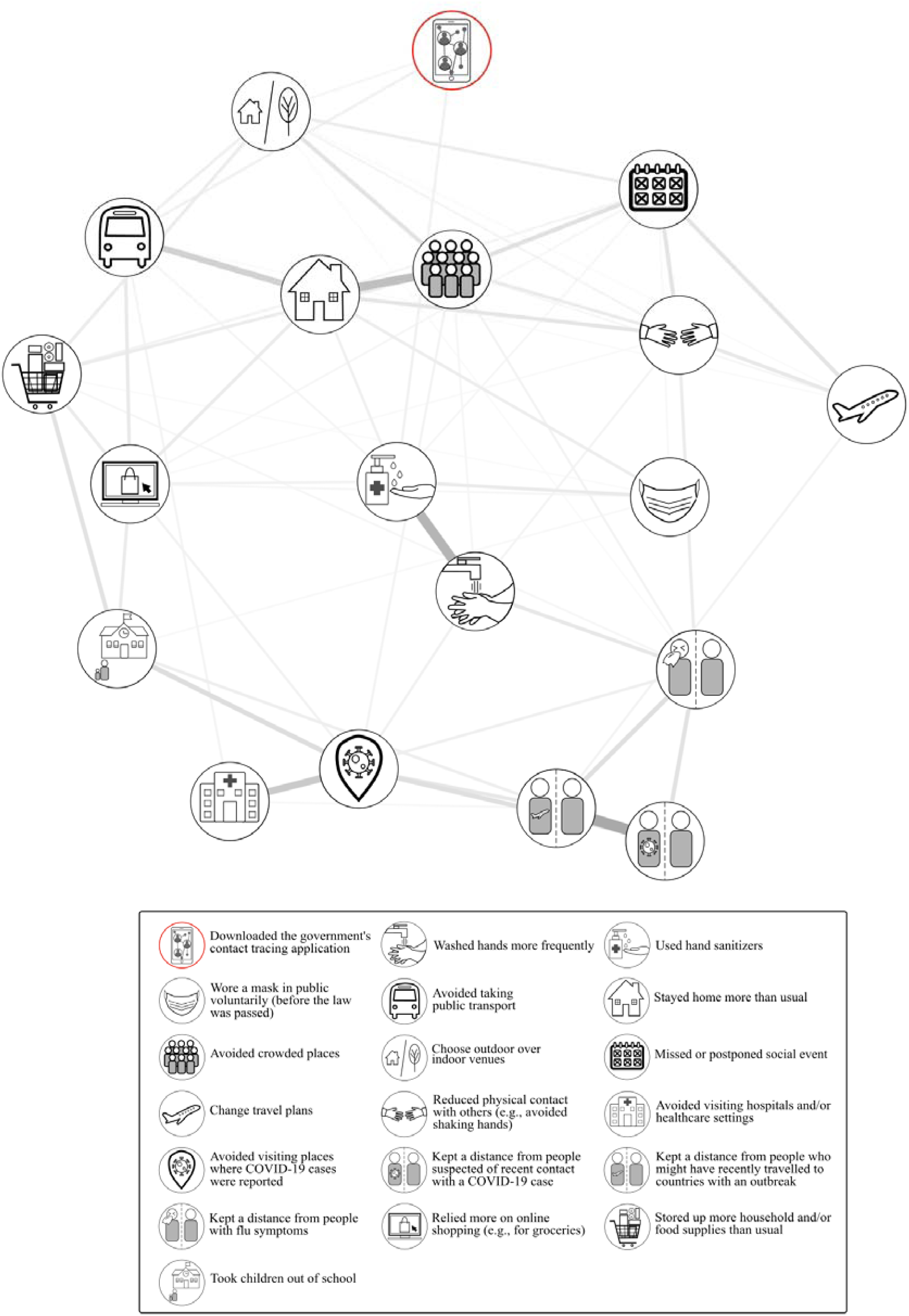
A model depicting how use of a contact tracing mobile application (TraceTogether) relates to other pandemic-related behavioral modifications. Here, line thickness represents the strength of an association.

(As a sensitivity analysis, we ran a logistic regression with use of the contract tracing application as the dependent variable, and the 18 other behavioral modifications as the predictors. Our conclusions did not change, see Appendix B.)

## 4. Discussion

As lockdowns for COVID-19 ease globally, digital contact tracing will play an increasingly critical role in managing the epidemic curve. However, this requires the public to actively download a contact tracing application – a step that has proven elusive to public health agencies worldwide [12]. Correspondingly, we identified for the first-time sociodemographic factors that predict voluntary use of a contact tracing application.

### 4.1 Behavioral Modifications

As our primary finding, we observed that the number of behavioral modifications significantly predicted the use of digital contact tracing. In other words, a person who had already changed his/her lifestyle on account of the pandemic was also likely to download a contact tracing application. Network analyses showed that downloads clustered with: (i) using hand sanitizers, (ii) avoiding public transport, and (iii) preferring outdoor over indoor venues. This may suggest that public health campaigns could capitalize on other behavioral modifications when seeking to promote application downloads – for example, by printing information about a contact tracing application on the packaging of hand sanitizers.

More broadly, our findings extend previous research on how individuals change their behaviors during a pandemic. Based on prior outbreaks, a taxonomy of modifications had been identified whereby: (i) ‘avoidant behaviors’ are measures taken to avoid contact with potential carriers (e.g., avoiding crowded places), while (ii) ‘prevention behaviors’ are those associated with maintaining hygiene (e.g., regular handwashing) [42]. Extrapolating to the technological realm, our findings suggest that using a contact tracing application cuts across this taxonomy, since downloads were linked to *both* avoidant (avoiding public transport, preferring outdoor venues) and prevention behaviors (using hand sanitizers). Moving forward, we thus urge further research to revise extant classification systems in light of technological innovations.

### 4.2 Demographic and Situational Factors

Aside from behavioral modifications, it is notable that no demographic (e.g., age, gender) or situational variable (e.g., number of COVID-19 cases, lockdown status) emerged as a predictor of digital contact tracing adoption. Prior to our study, it would have been conceivable that only a subset of the population would download a contact tracing application (e.g., demographic sectors based on gender, educational level, or age [26,40] [36]). By contrast, our findings highlight how take-up of digital contact tracing cuts across demographic groups.

As public health agencies develop strategies to promote downloads, this observation may suggest that demographic-specific messages are not needed. This is encouraging because the behavioral sciences offer widespread measures to ‘nudge’ the general population [41]. In this case, members of the public simply need a once-off nudge to download the application, following which the application functions independently in the background. Thus, if governments can nudge users in this first step (e.g., by introducing incentives to download, or by introducing contact tracing as an opt-out feature of existing government applications), it may be possible to attain the download rates necessary for contact tracing to be effective. At the same time, we urge further research into the acceptance of such strategies: given public concerns regarding privacy [24,25], any widespread interventions would need to be introduced cautiously.

### 4.3 Limitations

In discussing our findings, we note several limitations of our study. First, our survey relied on participants’ self-reported use of a contact tracing application. Although our download rate approximates that of the general population, further studies may seek to verify actual usage - for example, by incorporating survey questions into a contact tracing application. Second, we examined TraceTogether, a centralized contact tracing protocol. Future research will need to assess whether our findings extend to decentralized protocols, or to other forms of digital contact tracing that do not use phone applications (e.g., the public acceptance of cloud-based contact in South Korea).

### 4.4. Conclusion

To conclude, there is growing recognition that digital technology can contribute to pandemic management. What remains unclear, however, is how this technology is received and how best to promote uptake. Focusing on contact tracing, we found that downloads of a phone application was best predicted by the adoption of other infection control measures such as increased hand hygiene. In other words, introducing digital contact tracing is not merely a call to “TraceTogether” but to “modify together”, to use contact tracing applications as part of the broader spectrum of behavioral modifications during a pandemic.

## Data Availability

The data that support the findings of this study are available on request from the corresponding author (JCJL). The data are not publicly available due to IRB restrictions.

## Acknowledgements

This research was funded by a grant awarded to JCJL from the JY Pillay Global Asia Programme (grant number: IG20-SG002).

## Notes

### Competing Interest Statement

The authors have declared no competing interest.

### Clinical Trial

NCT04468581

### Author Declarations

Yale-NUS College Ethics Review Committee (#2020-CERC-001)

